# Risk factors of severity in community-acquired staphylococcal pneumonia

**DOI:** 10.1101/2020.07.31.20162875

**Authors:** Yves Gillet, Anne Tristan, Jean-Philippe Rasigade, Mitra Saadatian-Elahi, Coralie Bouchiat, Michele Bes, Oana Dumitrescu, Marie Leloire, Céline Dupieux, Frédéric Laurent, Gérard Lina, Jerome Etienne, Philippe Vanhems, Laurent Argaud, Francois Vandenesch, the PVL pneumonia study group

**Author notes:** Members listed in the Appendix. Corresponding author. Francois Vandenesch. Alternate corresponding author: Yves Gillet.

## Abstract

**Background:** *Staphylococcus aureus* causes severe forms of community-acquired pneumonia (CAP), namely staphylococcal pleuropneumonia in young children and staphylococcal necrotizing pneumonia in older patients. Methicillin resistance and the Panton-Valentine leukocidin (PVL) toxin have both been associated with poor outcome in severe CAP, but their respective roles are unclear.

**Methods:** Prospective multicenter cohort study of severe staphylococcal CAP conducted in 77 pediatric and adult intensive care units in France between January 2011 and December 2016. Clinical features and outcomes were compared between toddlers (<3 years of age) and older patients with PVL-positive CAP; and between older patients with PVL-negative or PVL-positive CAP. Risk factors for mortality were identified using multivariate Cox regression.

**Results:** Of 163 included patients, aged one month to 87 years, 85 (52.1%) had PVL-positive CAP; there were 20 (12.3%) toddlers, among whom 19 (95%) had PVL-positive CAP. The features of PVL-positive CAP in toddlers matched with the historical description of staphylococcal pleuropneumonia, with a lower mortality (n = 3/19, 15%) compared to PVL-positive CAP in older patients (n=31/66, 47%). Mortality in older patients was independently predicted by PVL-positivity (hazard ratio 1.81, 95% CI, 1.03 to 3.17) and methicillin resistance (2.37, 95% CI 1.29 to 4.34). As genetic diversity was comparably high in PVL-positive and PVL-negative isolates, confounding by microbial population structure was unlikely.

**Conclusion:** PVL was associated with staphylococcal pleuropneumonia in toddlers and was a risk factor for mortality in older patients with severe CAP, independently of methicillin resistance. Funded by the French ministry of Health (PHRC 2010-A01132-37).

## Introduction

Staphylococcal necrotizing pneumonia was first described in 1919 in adults during the influenza pandemic.^1^ It is associated with airway hemorrhage, epithelial necrosis, and a high fatality rate in otherwise healthy patients.^2^ It was seldom reported during the XX^th^ century^3,4^ until the epidemiological and pathophysiological links with Panton-Valentine Leucocidin (PVL) were made at the turn of the century.^2,5^ Subsequent reports confirmed the high fatality rates (40-50%) of PVL-associated pneumonia in adults.^6–9^

Independently of staphylococcal necrotizing pneumonia, staphylococcal pleuropneumonia (SPP) in young children was described during the late 1950s and 1960s as a specific clinical entity characterized by round-shaped lung infiltration evolving towards bullous lesions, purulent pleural effusion, and lack of airway hemorrhage or epithelial necrosis.^10,11^ Recent case series of SPP involving PVL-positive community-acquired *S. aureus*^12,13^ pointed to a possible relationship between SPP and PVL. In these series, the fatality rate was below 5%.These strikingly different diseases, both associated with PVL, may have contributed to controversies regarding the impact of PVL on the severity of staphylococcal pneumonia.^14–16^ In this context, whether age affects both the clinical picture and the outcome of community-acquired PVL-positive staphylococcal pneumonia, and whether PVL is an independent factor of severity are questions that remain to be answered and which are addressed herein.

## Patients and methods

### Ethics

The regional ethics committee approved the study (number: A11-39). Written informed consent was obtained from all patients or their parents.

### Study design and participants

Patients with *S. aureus* community-acquired pneumonia (CAP; criteria in Appendix) and admitted to a participating intensive care unit (ICU) were included between January 2011 and December 2016. Exclusion criteria were HIV-positive status, hospital-acquired pneumonia, or admission to hospital in the past 3 months. Clinical, laboratory, and therapeutic data were prospectively collected, and patients were assessed at admission to ICU, on day 1, 3, and 7. Severity was evaluated using age-adapted severity scores (Appendix).

### Microbiology

The causative staphylococcal strains were transmitted to the French National Reference Centre for Staphylococci for full characterization and microarray genotyping (Appendix).^17^

### Statistical analysis

We determined whether the previous descriptions of different PVL-associated diseases, namely SPP in children and staphylococcal necrotizing pneumonia in adults, correlated with identifiable age groups in patients with severe CAP. To this aim, patterns in patient age distribution were detected using a clustering procedure (Appendix, Fig. A1). Following this, disease presentation and outcome in toddlers (<3 years of age, as determined by the age clustering procedure) were compared to that found in older patients; and in older patients PVL-positive cases were compared to PVL-negative cases. The comparison of PVL-positive and PVL-negative cases was restricted to older patients (≥3 years of age) because 95% of toddlers had PVL-positive CAP. Between-group comparisons used Fisher’s exact test or Student’s t-test, as appropriate. Possibly non-linear changes of mortality rates in function of patient age were visualized using kernel density estimation with bootstrap confidence intervals (Appendix). Potential predictors of mortality were investigated using univariate and multivariate Cox proportional hazards models. To avoid bias due to row-wise deletion of cases containing missing data in multivariate models,^18^ all missing data were imputed prior to analysis using a non-linear, random forest-based multiple imputation technique. Imputation uncertainty was accounted for by replacing binary factors (such as, for example, leukopenia) by a probability estimate between 0 (absence) and 1 (presence). Analyses were conducted using R software v3.6.0 (R Core Team [2019]. R: A language and environment for statistical computing. R Foundation for Statistical Computing, Vienna, Austria. URL https://www.R-project.org/). Anonymized data and software code required to reproduce the results are available at github.com/rasigadelab/severecap.

## Results

### Clustering analysis of patient age identifies specific patterns in PVL-positive and PVL-negative *S. aureus* pneumonia

A total of 245 patients were identified, and after removal of those who did not comply with the inclusion criteria, without consent, and those with missing CRF, 163 patients were included (Appendix, Fig. A1). These were hospitalized in 77 ICUs, and aged from one month to 87 years. Overall, 85 patients (52.1%) were infected by a PVL-positive *S. aureus*. Clustering of patients with PVL-positive CAP identified two non-overlapping clusters below and above ∼three years of age, while the analysis of PVL-negative *S. aureus* patients identified two overlapping clusters centered at 28 and 60 years of age (Appendix, Fig. A2). Thus, almost all cases in patients aged of <3 years (hereinafter “toddlers”) were PVL-positive while, in older patients, PVL positivity was evenly distributed according to age. In contrast, PVL-negative CAP was exceptional in younger children (only one case under 15 years of age) and its prevalence increased with patient age, peaking at 60 years. The microbiological and clinical features of CAP differed markedly between toddlers (n = 20) and older patients (n = 143). PVL positivity was more frequent in toddlers (n = 19/20 vs. 77/143; odds ratio 21.9, 95% CI 3.3 to 928.6), more strongly so than methicillin resistance (n = 7/20 vs. 23/143, odds ratio 2.8, 95% CI 0.85 to 8.6). Additional results concerning the specificities observed in toddlers are developed in the Appendix.

### Genetic diversity of causative isolates

Strain genetic background were extremely diverse; there were 13 different clonal complexes (CC) in PVL-positive and 17 CC in PVL-negative isolates, 7 of those CCs being shared by the two groups (further details in Appendix).

### Mortality according to age

Mortality peaked at ∼30 years and above 60 years (Fig. 1a-b). The mortality peak at ∼30 years was more pronounced in PVL-positive cases (Fig. 1c-d) while the mortality peak above 60 years was stronger in PVL-negative cases (Fig. 1e-f). Mortality was lower in toddlers than older patients (n = 3/20 vs. 52/143, odds ratio 0.31, 95% CI 0.06 to 1.15). In toddlers with PVL-positive CAP (Fig. 1 g-h), most cases occurred before 12 months of age and were non-lethal, while mortality increased after one year of age. Since only 1 toddler had PVL-negative CAP, we could not compare the outcomes of PVL-positive to PVL-negative CAP in this group. Further analysis aiming to compare PVL-positive to PVL-negative cases was restricted to patients ≥3 years to avoid the interpretation bias that would have resulted from the pooling of markedly different patients (namely, toddlers and older patients) in a single analysis.

**Figure 1:**
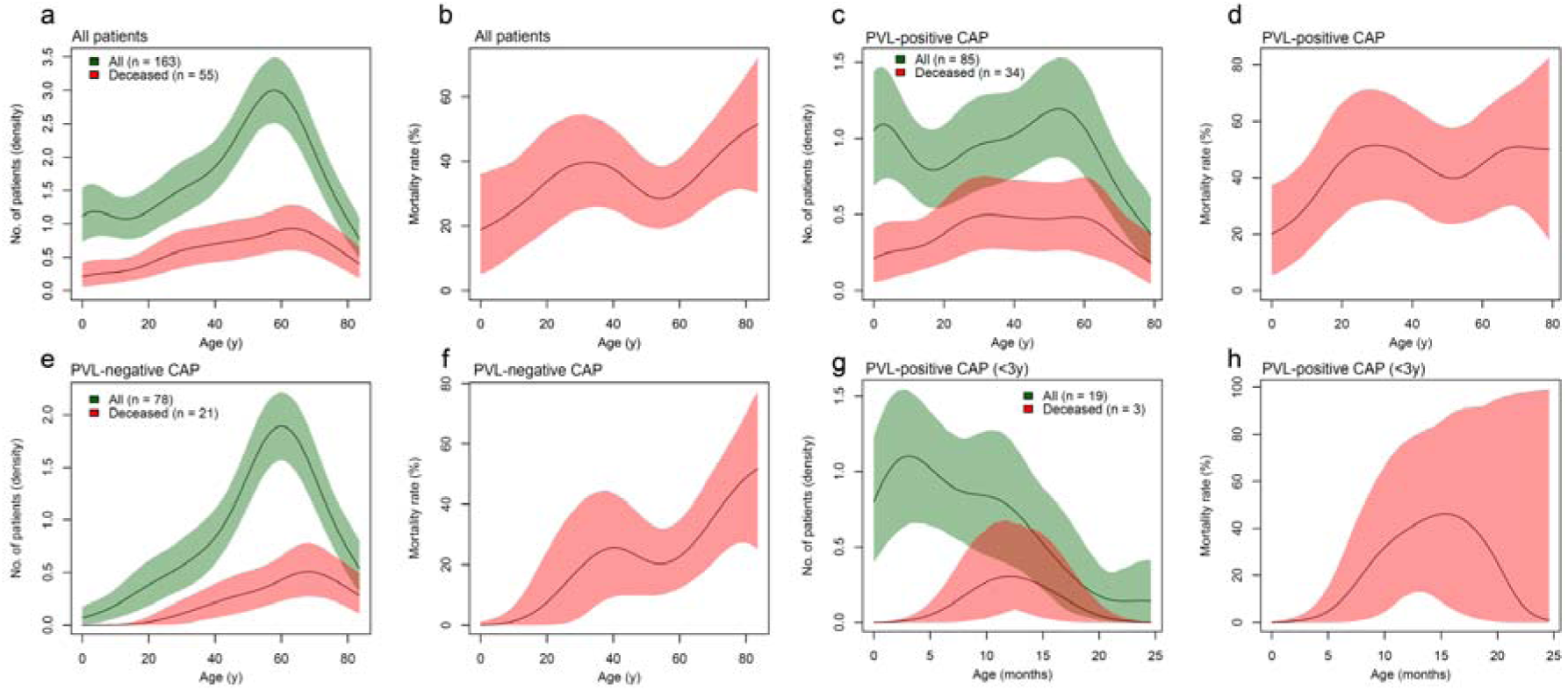
Mortality of patients with *S. aureus* community-acquired pneumonia (CAP) according to age and the presence of the Panton-Valentine leukocidin (PVL). Panels a, c, e and g present density estimates of the occurrence of pneumonia in all patients of the considered group (green lines) and in deceased patients (red lines). Panels b, d, f, and h present estimates of mortality rate. Colored areas present bootstrap-based 95% confidence bands of the estimates. Analyses were conducted separately for all patients (a and b), patients with PVL-positive CAP (c and d), patients with PVL-negative CAP (e and f), and patients <3 years of age (toddlers) with PVL-positive CAP (g and h).

### PVL is a severity factor in patients ≥3 years

Among the 143 patients ≥3 years, 66 (46.2%) had PVL-positive CAP. Compared to PVL-negative CAP, patients with PVL-positive CAP were younger and less likely to have an underlying condition (Table 1). At admission, they had more frequently airway hemorrhage and cutaneous eruption or rash evoking immune system reaction. PVL-positive CAP patients were also more likely to present with septic shock, leukopenia, elevated blood lactate and SOFA severity score, and to require extra corporeal membrane oxygenation (ECMO).

**Table 1:**
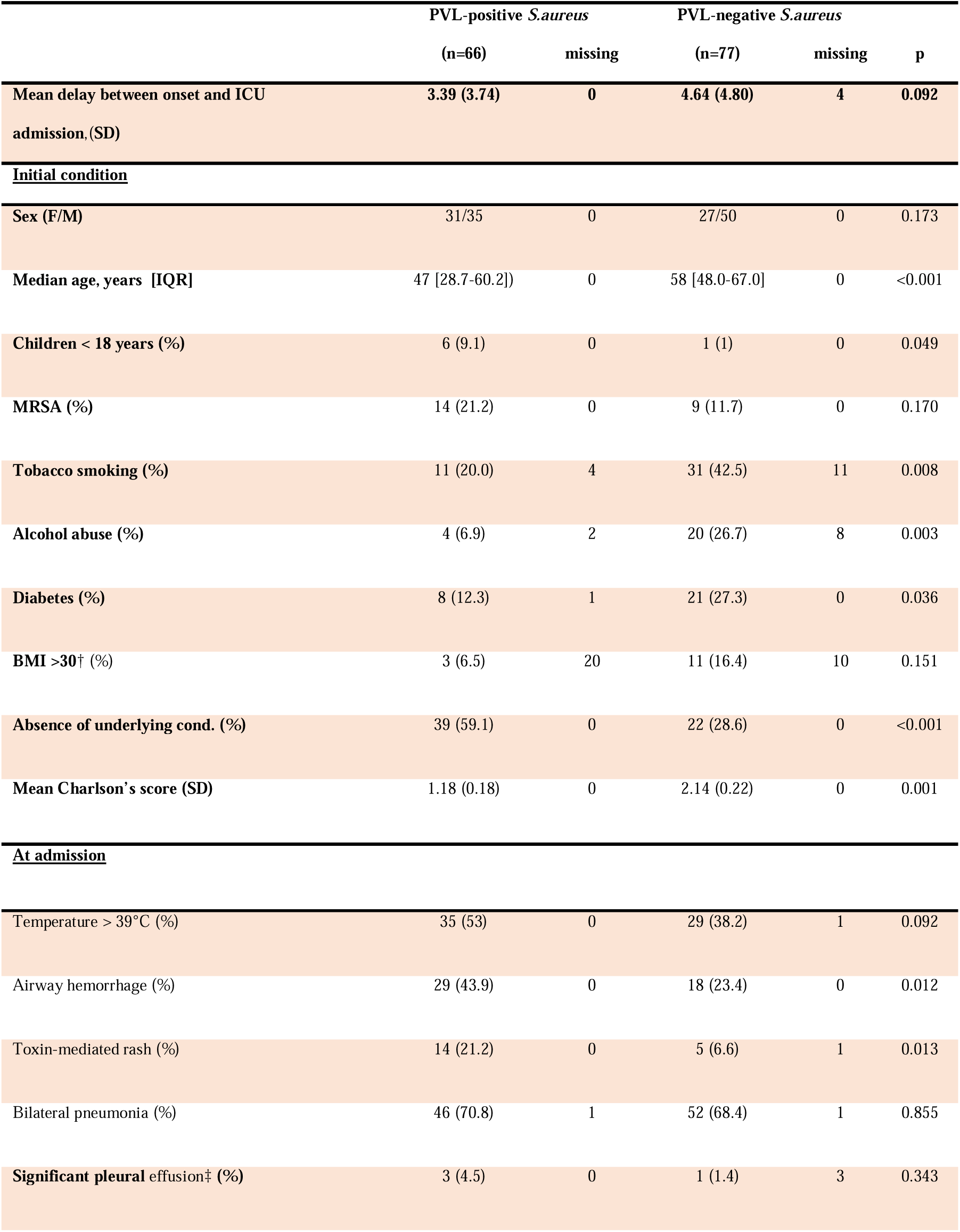
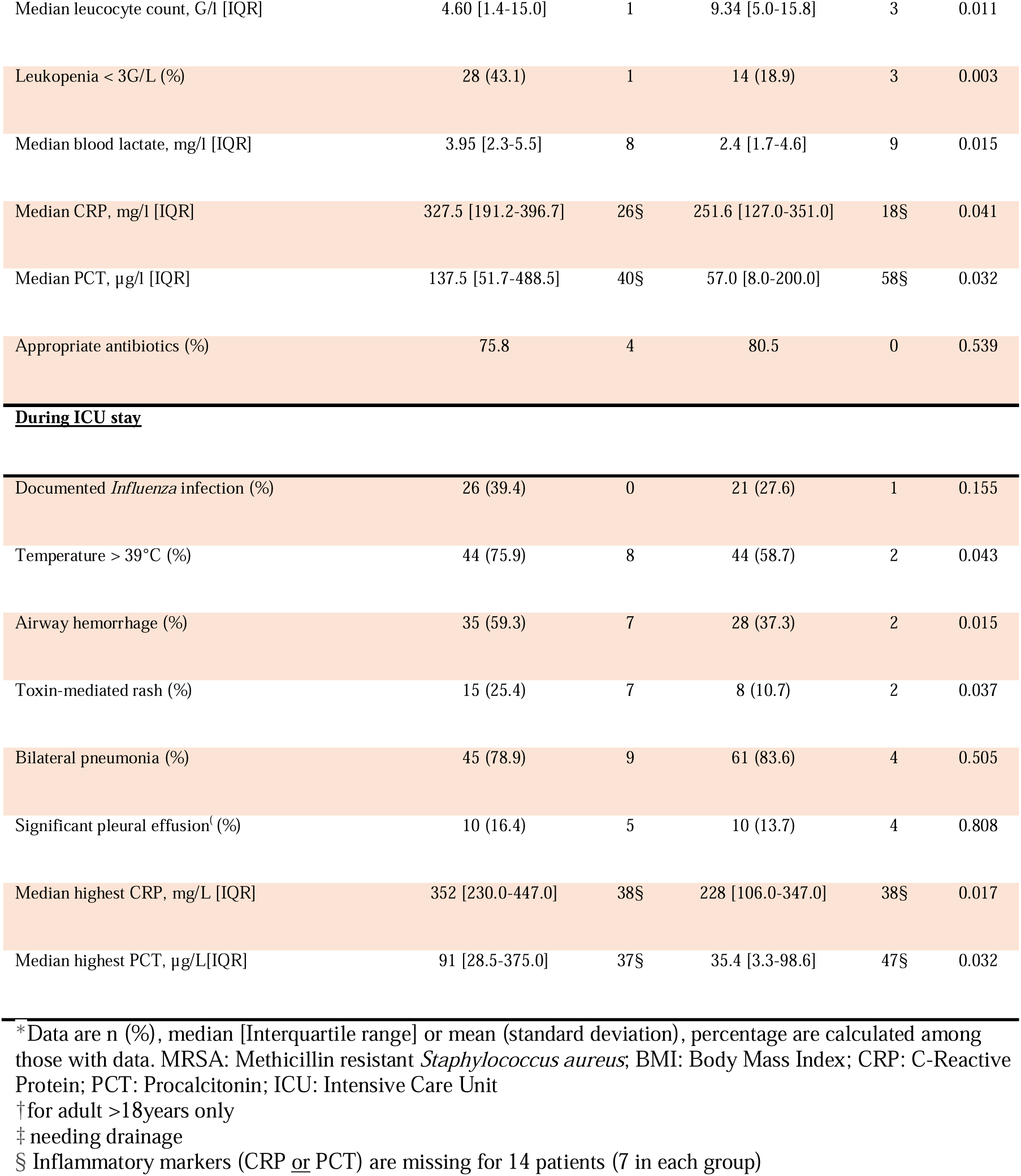
Clinical and laboratory characteristics at admission and during stay according to PVL status in patients ≥3 years of age.*

Appropriate initial antibiotic treatment, defined as at least one drug active against the corresponding strain, was equally likely in PVL-positive and PVL-negative CAP. However, treatment was less frequently appropriate in MRSA cases (odds ratio 0.027, 95% CI 0.006 to 0.097). See Appendix for additional information on antibiotic treatment.

Most of the differences observed between patients with PVL-positive and PVL-negative CAP at admission persisted during the first week in ICU (Table 2). The higher SOFA score at admission persisted at day 1, as did the higher median blood lactate. Temperature >39°C, airway hemorrhage, and rash remained significantly more common in PVL-positive cases, who received more frequently inhaled nitric oxide and required ECMO support.

**Table 2:** Severity scores and markers at admission and during evolution according to PVL status in patients ≥3 years of age.

|  | PVL-positive *S.aureus* |  | PVL-negative *S.aureus* |  |  |
| --- | --- | --- | --- | --- | --- |
|  | n=66 | missing | n=77 | missing | p |
| Admission |  |  |  |  |  |
| Mean SOFA score (SD) | 8.9 (5.68) | 2 | 6.9 (5.01) | 8 | 0.004 |
| Septic shock (%) | 31 (48.4) | 4 | 22 (30.1) | 2 | 0.035 |
| ARDS (%) | 20 (32.8) | 7 | 16 (22.9) | 5 | 0.352 |
| Severe ARDS (%) | 12 (19.7) | 7 | 10 (14.3) | 5 | 0.485 |
| Invasive ventilation (%) | 32 (48.5) | 0 | 31 (42.5) | 4 | 0.499 |
| ECMO (%) | 11 (16.9) | 1 | 3 (4.2) | 5 | 0.022 |
| Day 1 |  |  |  |  |  |
| Mean SOFA score (SD) | 10.9 (5.96) | 14 | 8.8 (5.25) | 8 | 0.040 |
| Septic shock (%) | 31 (54.4) | 9 | 28 (40.6) | 8 | 0.152 |
| ARDS (%) | 27 (50) | 12 | 32 (50) | 13 | 1 |
| Severe ARDS (%) | 21 (38.9) | 12 | 19 (29.7) | 13 | 0.332 |
| Death (%) | 7 (10.6) | 0 | 4 (5.2) | 0 | <0.001 |
| Mean variation in blood lactate<sup>†</sup>, mg/l (SD) | 0.62 (4.38) | 19 | - 0.69 (2.10) | 14 | 0.063 |
| Day 3 |  |  |  |  |  |
| Mean SOFA score (SD) | 10.6 (6.51) | 24 | 8.1 (5.34) | 8 | 0.045 |
| Cumulative death (%) | 17 (25.8) | 0 | 7 (9.1) | 0 | <0.001 |
| Day 7 |  |  |  |  |  |
| Mean SOFA score (SD) | 8 (6.84) | 29 | 6.8 (5.70) | 16 | 0.347 |
| Cumulative death (%) | 23 (34.8) | 0 | 10 (13) | 0 | <0.001 |
| During whole ICU stay |  |  |  |  |  |
| Invasive ventilation (%) | 56 (84.8) | 0 | 60 (81.1) | 3 | 0.655 |
| Inhaled NO (%) | 21 (33.3) | 3 | 11 (15.3) | 5 | 0.016 |
| ECMO (%) | 20 (32.3) | 4 | 8 (11) | 4 | 0.003 |
| Death (%) | 31 (47.0) | 0 | 21 (27.3) | 0 | 0.023 |

\* Data are n (%), mean (standard deviation), percentage are calculated among those with data. SOFA: Sequential Organ Failure Assessment; ARDS: Acute Respiratory Distress Syndrome; NO: Nitrogen Monoxide; ECMO: Extra Corporeal Membrane Oxygenation
<sup>†</sup>between Day 0 and Day one

### Survival

We used survival analysis to examine the influence of microbiological and clinical factors on lethal outcome in the 143 patients ≥3 years of age. Kaplan-Meier survival curves suggested that PVL-positivity and methicillin resistance contributed additively to mortality (Fig. 2); median survival time was 1 day (interquartile range, IQR 0 to 7 days) for PVL-positive cases and 7 days (IQR 3 to 14 days; P = 0.02, Mann-Whitney *U*-test) for PVL-negative cases. Among the 91 survivors, length of ICU stay was longer in those with PVL-positive CAP (median time until discharge, 39 days, IQR 26 to 68 days) than in those with PVL-negative CAP (29 days, IQR 20 to 42 days; P = 0.01, Mann-Whitney *U*-test). A multivariate Cox regression model also supported an independent association of PVL-positivity (adjusted hazard ratio, aHR 1.8, 95% CI 1.04 to 3.2) and methicillin-resistance (aHR 2.4, 95% CI 1.3 to 4.4) with mortality. We used bivariate and multivariate Cox proportional hazards models to identify potential independent risk factors for mortality (Table 3). Predictors were included in the models based on their expected relevance to disease outcome. These included baseline patient characteristics, namely, sex and the Charlson comorbidity score; microbiological factors, namely, PVL and methicillin resistance; patient characteristics reflecting: (1) severity upon admission, namely, the SOFA score, hemoptysis, leukopenia and blood lactates, (2) inflammatory reaction, namely rash and blood procalcitonin, (3) a risk factor for staphylococcal superinfection of the lung, namely, a flu-like illness; and the appropriateness and expected toxin-suppressing activity of the antibiotics received (see Appendix for details).

**Table 3:** Cox regression analysis of predictors of death in patients ≥3 years with *S. aureus* pneumonia, including clinical predictors at admission and microbiological predictors.

| Predictor | Hazard ratio (95% confidence interval) |  |  |
| --- | --- | --- | --- |
|  | Bivariate models | Full model | Best-fitting model |
| Charlson comorbidity score (per point) | 1.05 (0.91 to 1.21) | 1.03 (0.85 to 1.26) | - |
| Male sex | 0.68 (0.39 to 1.17) | 0.95 (0.51 to 1.78) | - |
| PVL | 1.95 (1.12 to 3.41) | 0.87 (0.42 to 1.83) | - |
| Methicillin resistance | 2.57 (1.41 to 4.69) | 3.86 (1.34 to 11.16) | 2.87 (1.53 to 5.40) |
| SOFA score | 1.11 (1.05 to 1.16) | 1.03 (0.96 to 1.11) | - |
| Flu-like illness | 0.71 (0.41 to 1.23) | 0.32 (0.15 to 0.68) | 0.32 (0.17 to 0.60) |
| Hemoptysis | 2.62 (1.52 to 4.52) | 1.87 (0.98 to 3.54) | 2.15 (1.17 to 3.93) |
| Rash | 2.17 (1.12 to 4.23) | 2.59 (1.22 to 5.48) | 2.52 (1.24 to 5.12) |
| Leukopenia | 3.45 (2.00 to 5.97) | 2.38 (1.19 to 4.76) | 2.38 (1.23 to 4.58) |
| Blood procalcitonin (per 2-fold increase) | 1.40 (1.21 to 1.63) | 1.05 (0.88 to 1.24) | - |
| Blood lactates (per 2-fold increase) | 2.40 (1.85 to 3.12) | 2.23 (1.52 to 3.28) | 2.64 (1.95 to 3.57) |
| Adapted antimicrobial therapy | 0.82 (0.44 to 1.53) | 1.54 (0.52 to 4.59) | - |
| Antitoxin therapy | 2.67 (1.48 to 4.81) | 1.29 (0.63 to 2.65) | - |

NOTE. Bivariate models were independent Cox proportional hazards models, one per predictor. The full multivariate model included all predictors (likelihood ratio test, $P < 0.001$ ; Akaike information criterion [AIC], 420 with 13 degrees of freedom). The best-fitting model was obtained using a stepwise procedure, starting from the full model, and minimizing the AIC (409 with 6 degrees of freedom). Confidence interval widths were not corrected for test multiplicity. PVL, Panton-Valentine leukocidin. SOFA score, sequential organ failure assessment score.

**Figure 2:**
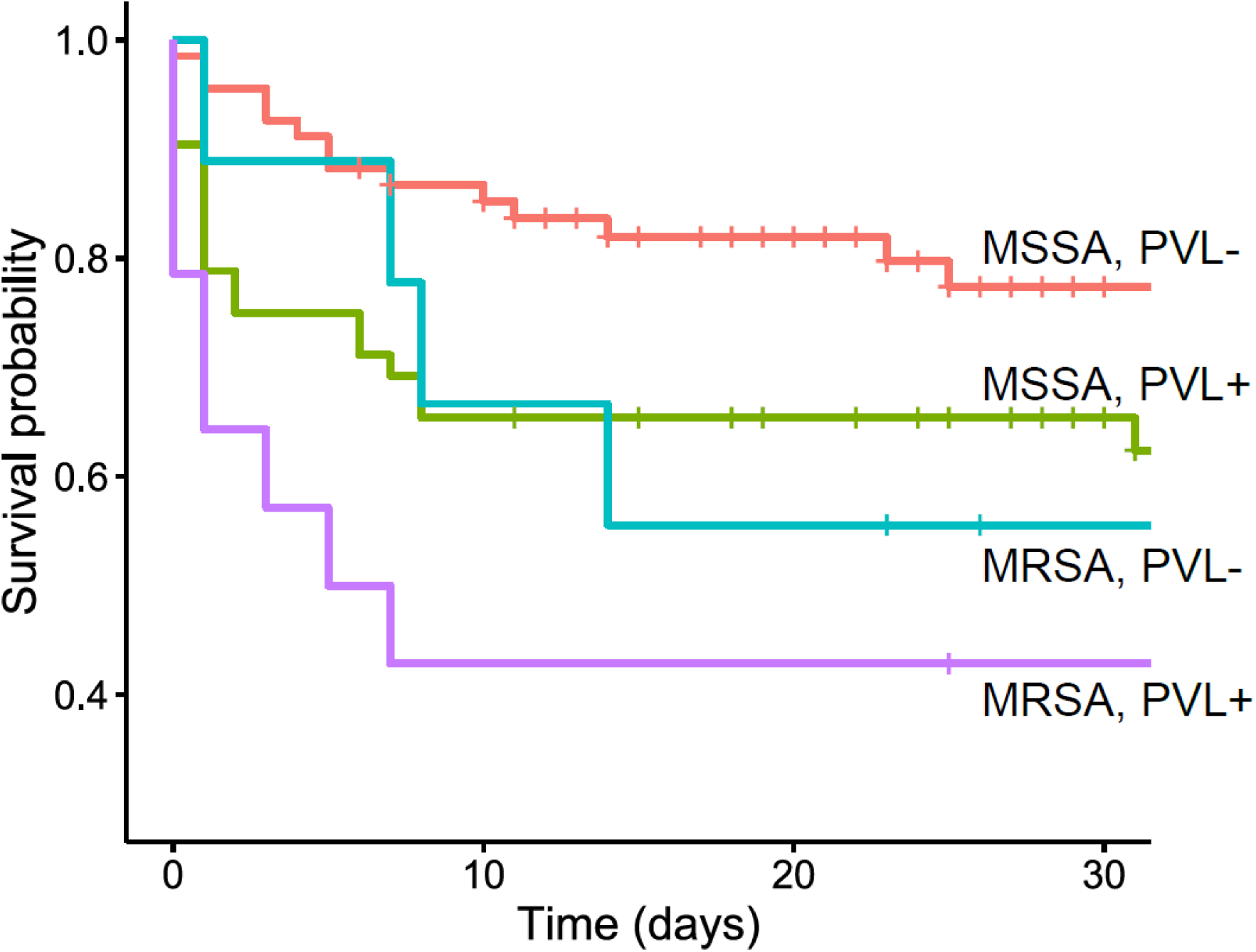
Survival of 143 patients ≥3 years of age with *S. aureus* pneumonia. Survival after ICU admission according to whether the causative strain was methicillin-resistant (MRSA) or -susceptible (MSSA) and whether it harbored the PVL toxin are shown. ‘+’ marks denote censored (discharged) patients. Log-rank test, p=0.0007.

In the best-fitting multivariate model, the independent predictors of death were methicillin resistance, hemoptysis, rash, leukopenia, elevated blood lactates, and the absence of a flu-like illness. Interestingly, the best-fitting model did not include PVL-positivity but included previously recognized severity factors in PVL-positive lung infection,^19^ namely, hemoptysis and leukopenia. The exclusion of PVL, but not its clinical consequences, from the final model is suggestive of a chain of consequence between PVL, severity factors and death, where the severity factors mediate the effect of PVL on death.^20^ See Appendix for additional data on severity-associated features and on interactions between predictors of mortality.

## Discussion

As the present study is one of the largest prospective series of PVL-positive *S. aureus* pneumonia and is the first to compare PVL-positive and -negative cases since the description of staphylococcal necrotizing pneumonia in 2002,^2^ we are able to better understand the role of PVL in different clinical presentation and in severity of staphylococcal pneumonia.

PVL-positive CAP and its severity was not evenly distributed according to age. PVL-negative CAP was virtually absent in toddlers and mortality was low. Many infectious processes vary in terms of presentation between young and older subjects, but the differences we observed exceed the expected variations due to age alone. For instance, radiological findings in pneumonia are usually similar at all ages, ^21^ however, we observed substantial radiological differences, notably regarding pleural effusion, pneumothorax and unilateral involvement that were more frequent in toddlers. It was also unexpected to observe a lower mortality in very young children compared to young adults without underlying conditions. Thus, despite similarities in terms of site of infection and bacteriology, staphylococcal CAP of the young children (i.e. SPP) and staphylococcal necrotizing pneumonia of the adults should be considered as two distinct entities.

As we uncovered major differences according to age in PVL-positive staphylococcal pneumonia, the lack of consideration of age-specificity of the disease severity may at least partially explain some conflicting results reported in the literature.^14,22^ Considering that PVL-negative staphylococcal pneumonia are almost absent in the youngest patients, toddlers should be excluded from analysis to assess the role of PVL in severity.

We observed that PVL was associated with specific symptoms in staphylococcal pneumonia of adolescents and adults. Previously described features strongly associated with PVL^2,7^ were observed herein among patients aged ≥3 years: PVL-positive *S. aureus* pneumonia occurred mainly in younger people without underlying condition and was associated with cutaneous rash, airway hemorrhage, and leukopenia. Strikingly, PVL was associated with rash independently of major superantigens. Because PVL itself has no known superantigenic properties, it is unclear whether this association reflects a direct effect of PVL or a less-specific consequence of severe inflammation.

Airway hemorrhage is a consequence of respiratory epithelial necrosis, as described in lung autopsies,^1,2^ and is an indirect effect of PVL since the presence of polymorphonuclear (PMN) cells in the lung is required for PVL-induced necrotic lesions of the epithelium.^23^ PMN cell influx to the lung may be generated by two non-exclusive mechanisms: a preceding viral infection producing chemoattractant for neutrophils by epithelial cells,^23^ and a consequence of PVL-mediated inflammasome activation, leading to PMN cell recruitment and activation.^24^ One objective of the present study was to assess the link between PVL and severity, considering that such an association has been controversial since the first description of PVL-associated pneumonia in 2002. ^2^ The frequency of PVL in severe CAP in adults herein contrasts with the low frequency of PVL (<5%) in the staphylococcal carriage population in France^17^ and this striking difference constitutes the initial evidence for an association of PVL with severity. We observed that PVL, especially but not exclusively when associated with methicillin resistance, was an independent factor of mortality in staphylococcal CAP, outweighing protective factors such as young age and absence of underlying conditions. The present study indicates that PVL is associated with non-specific severity markers of infection such as elevated lactate or presence of septic shock at admission and absence of reduction in lactate between admission and day 1. All these factors are associated with a higher mortality in sepsis, irrespective of the bacteria involved,^25,26^ and their higher frequency in the PVL-positive cases confirms greater severity in such patients. Surrogate markers of respiratory failure, such as the need for nitric oxide and for ECMO, were more frequent in PVL-positive cases, which also indicates greater severity in accordance with recent studies.^27-28^ The symptoms associated with PVL (i.e. rash, airway hemorrhage, and leukopenia) were associated with mortality in the multivariate prediction model, strongly suggesting that these clinical factors mediate the link between PVL and death.

Since its first description,^2^ PVL-positive *S. aureus* pneumonia has been reported worldwide.^28–31^ A particular situation is observed in the USA where the description of necrotizing pneumonia coincided with the emergence of the PVL-positive CA-MRSA USA300 lineage.^4,32^ PVL has been also associated with the CA-MRSA ST-80 lineage in Europe.^33^ However, only 11 and five strains of the present study belong to these clones, respectively, and PVL was distributed within a large diversity of genetic background (i.e. 13 CCs) with a majority of MSSA, ruling out a possible clonal bias associated with PVL-positive cases. The particular lineage restriction observed in the US may have led to confusion in the understanding of the determinant of severity associated with staphylococcal CAP. Thus, methicillin resistance was thought to be the prominent determinant for severity, presumably by inducing a delay in initiation of appropriate antibiotics. The present study found that patients infected with MRSA were less likely to have received appropriate antibiotics at admission and had a worse prognosis. However, this higher mortality in MRSA was present both in PVL-positive and PVL-negative-pneumonia patients.

The present study does, however, have certain limitations, most notably the observational design whereby all participating centers were encouraged to enroll patients fulfilling the inclusion criteria. We cannot exclude under-reporting, and a higher reporting of the most severe cases may have occurred, but this is unlikely to have affected directly the comparison between PVL-positive and PVL-negative cases. The number of missing data were limited and exhaustivity was reached for all major parameters. Furthermore, the study was restricted to France but covers almost entirely the French territory with 77 participating centers thus limiting possible local epidemiological bias.

In conclusion, the present study demonstrates the association of PVL with two distinct facets of staphylococcal CAP with marked differences between toddlers and adolescents/adults regarding clinical presentation and outcome. In toddlers, PVL appears to be prominent in staphylococcal pneumonia, presentation matches with SPP, and standard of care in modern ICU appears to be sufficient for favorable outcome. In contrast, PVL-positive CAP in adolescents and adults remains extremely severe despite aggressive management; it deserves further research to develop a novel therapeutic approach.

## Data Availability

not an interventional study

https://github.com/rasigadelab/severecap

## Contributors

YG designed the study, analyzed and interpreted data, and wrote the report

AT collected and analyzed data, and revised the report.

JPR performed statistical analysis, interpreted data and wrote the report.

MSE contributed to design the study, collected data, and revised the report.

CB collected and analyzed data, and revised the report.

MB analyzed the microbiological data, and revised the report.

OD collected and analyzed data, and revised the report.

ML collected and analyzed data, and revised the report.

CD collected and analyzed data, and revised the report.

FL analyzed the microbiological data, and revised the report.

GL contributed to design the study and revised the report.

JE contributed to design the study and wrote the report.

PV contributed to design the study, contributed to statistical analysis, interpreted data and revised the report.

LA designed the study, interpreted data, and revised the report.

FV designed the study, acquired funding and ethics approval, is the chief investigator of the study, and wrote the report.

All authors read and approved the final report.

## Acknowledgments

The study was funded by French Ministry of Health under the Programme Hospitalier de Recherche Clinique 2011. This report presents independent research commissioned by the Ministry of Health; the views and opinions expressed in this publication are those of the authors and do not necessarily reflect those of the Ministry of Health. We thank the technicians of the French Reference Centre for Staphylococci for their contribution in characterizing *S. aureus* isolates. Finally, we thank Philip Robinson (DRCI, Hospices Civils de Lyon) for copy-editing the manuscript.

## References

1 Chickering HT, Park JHJ. Staphylococcus aureus pneumonia. JAMA 1919;72:617–26.

2 Gillet Y, Issartel B, Vanhems P, et al. Association between Staphylococcus aureus strains carrying gene for Panton-Valentine leukocidin and highly lethal necrotising pneumonia in young immunocompetent patients. Lancet 2002;359:753–9.

3 Wollenman OJ, Finland M. Pathology of Staphylococcal Pneumonia Complicating Clinical Influenza. Am J Pathol 1943;19:23–41.

4 Centers for Disease Control and Prevention (CDC). Four pediatric deaths from community-acquired methicillin-resistant Staphylococcus aureus — Minnesota and North Dakota, 1997-1999. MMWR Morb Mortal Wkly Rep 1999;48:707–10.

5 Lina G, Piémont Y, Godail-Gamot F, et al. Involvement of Panton-Valentine leukocidin-producing Staphylococcus aureus in primary skin infections and pneumonia. Clin Infect Dis 1999;29:1128–32.

6 David MZ, Daum RS. Community-associated methicillin-resistant Staphylococcus aureus: epidemiology and clinical consequences of an emerging epidemic. Clin Microbiol Rev 2010;23:616–87.

7 Kreienbuehl L, Charbonney E, Eggimann P. Community-acquired necrotizing pneumonia due to methicillin-sensitive Staphylococcus aureus secreting Panton-Valentine leukocidin: a review of case reports. Ann Intensive Care 2011;1:52.

8 Li H-T, Zhang T-T, Huang J, Zhou Y-Q, Zhu J-X, Wu B-Q. Factors associated with the outcome of life-threatening necrotizing pneumonia due to community-acquired Staphylococcus aureus in adult and adolescent patients. Respiration 2011;81:448–60.

9 Mandell LA, Wunderink R. Methicillin-resistant Staphylococcus aureus and community-acquired pneumonia: an evolving relationship. Clin Infect Dis 2012;54:1134–6.

10 Hendren WH, Haggerty RJ. Staphylococcic pneumonia in infancy and childhood; analysis of seventy-five cases. J Am Med Assoc 1958;168:6–16.

11 Rebhan AW, Edwards HE. Staphylococcal pneumonia: a review of 329 cases. Can Med Assoc J 1960;82:513–7.

12 Carrillo-Marquez MA, Hulten KG, Hammerman W, Lamberth L, Mason EO, Kaplan SL. Staphylococcus aureus pneumonia in children in the era of community-acquired methicillin-resistance at Texas Children’s Hospital. Pediatr Infect Dis J 2011;30:545–50.

13 Lemaître C, Angoulvant F, Gabor F, et al. Necrotizing Pneumonia in Children: Report of 41 Cases Between 2006 and 2011 in a French Tertiary Care Center. Pediatr Infect Dis J 2013;32:1146–9.

14 Shallcross LJ, Fragaszy E, Johnson AM, Hayward AC. The role of the Panton-Valentine leucocidin toxin in staphylococcal disease: a systematic review and meta-analysis. Lancet Infect Dis 2013;13:43–54.

15 Bubeck Wardenburg J, Palazzolo-Ballance AM, Otto M, Schneewind O, DeLeo FR. Panton-Valentine leukocidin is not a virulence determinant in murine models of community-associated methicillin-resistant Staphylococcus aureus disease. J Infect Dis 2008;198:1166– 70.

16 Diep BA, Gillet Y, Etienne J, Lina G, Vandenesch F. Panton-Valentine leucocidin and pneumonia. Lancet Infect Dis 2013;13:566.

17 Tristan A, Rasigade J-P, Ruizendaal E, et al. Rise of CC398 lineage of Staphylococcus aureus among Infective endocarditis isolates revealed by two consecutive population-based studies in France. PLoS One 2012;7:e51172.

18 van der Heijden Gjmg, Donders ART, Stijnen T, Moons KGM. Imputation of missing values is superior to complete case analysis and the missing-indicator method in multivariable diagnostic research: a clinical example. J Clin Epidemiol 2006;59:1102–9.

19 Gillet Y, Vanhems P, Lina G, et al. Factors predicting mortality in necrotizing community-acquired pneumonia caused by Staphylococcus aureus containing Panton-Valentine leukocidin. Clin Infect Dis 2007;45:315–21.

20 Richiardi L, Bellocco R, Zugna D. Mediation analysis in epidemiology: methods, interpretation and bias. Int J Epidemiol 2013;42:1511–9.

21 Patterson CM, Loebinger MR. Community acquired pneumonia: assessment and treatment. Clin Med (Lond) 2012;12:283–6.

22 Diep BA, Palazzolo-Ballance AM, Tattevin P, et al. Contribution of Panton-Valentine leukocidin in community-associated methicillin-resistant Staphylococcus aureus pathogenesis. PLoS One 2008;3:e3198.

23 Niemann S, Ehrhardt C, Medina E, et al. Combined action of influenza virus and Staphylococcus aureus panton-valentine leukocidin provokes severe lung epithelium damage. J Infect Dis 2012;206:1138–48.

24 Perret M, Badiou C, Lina G, et al. Cross-talk between Staphylococcus aureus leukocidins-intoxicated macrophages and lung epithelial cells triggers chemokine secretion in an inflammasome-dependent manner: Macrophage-epithelia cross-talk upon PVL treatment. Cell Microbiol 2012;14:1019–36.

25 Singer M, Deutschman CS, Seymour CW, et al. The Third International Consensus Definitions for Sepsis and Septic Shock (Sepsis-3). JAMA 2016;315:801–10.

26 Casserly B, Phillips GS, Schorr C, et al. Lactate measurements in sepsis-induced tissue hypoperfusion: results from the Surviving Sepsis Campaign database. Crit Care Med 2015;43:567–73.

27 Gijón M, Bellusci M, Petraitiene B, et al. Factors associated with severity in invasive community-acquired Staphylococcus aureus infections in children: a prospective European multicentre study. Clin Microbiol Infect 2016;22:643.e1-6.

28 Jacquot A, Luyt C-E, Kimmoun A, Levy B, Baux E, Fluvalentine Study group. Epidemiology of post-influenza bacterial pneumonia due to Panton-Valentine leucocidin positive Staphylococcus aureus in intensive care units: a retrospective nationwide study. Intensive Care Med 2019;45:1312–1314.

29 Hsu L-Y, Koh T-H, Anantham D, Kurup A, Chan KPW, Tan B-H. Panton-Valentine leukocidin-positive Staphylococcus aureus, Singapore. Emerg Infect Dis 2004;10:1509–10.

30 Holmes A, Ganner M, McGuane S, Pitt TL, Cookson BD, Kearns AM. Staphylococcus aureus isolates carrying Panton-Valentine leucocidin genes in England and Wales: frequency, characterization, and association with clinical disease. J Clin Microbiol 2005;43:2384–90.

31 Francis JS, Doherty MC, Lopatin U, et al. Severe community-onset pneumonia in healthy adults caused by methicillin-resistant Staphylococcus aureus carrying the Panton-Valentine leukocidin genes. Clin Infect Dis 2005;40:100–7.

32 Finelli L, Fiore A, Dhara R, et al. Influenza-associated pediatric mortality in the United States: increase of Staphylococcus aureus coinfection. Pediatrics 2008;122:805–11.

33 Dufour P, Gillet Y, Bes M, et al. Community-acquired methicillin-resistant Staphylococcus aureus infections in France: emergence of a single clone that produces Panton-Valentine leukocidin. Clin Infect Dis 2002;35:819–24.

